# Effect of Ambient Temperature on Diabetes Patient Visits at Primary and Referral Healthcare Services in Yogyakarta Province, Indonesia: An Analysis of Indonesian National Health Insurance Data 2021

**DOI:** 10.1101/2023.08.16.23291838

**Authors:** Aditya Lia Ramadona, Hayu Qaimamunazzala, Hari Kusnanto, Lutfan Lazuardi, Cornelia Wahyu Danawati, Anis Fuad, Fatwa Sari Tetra Dewi

## Abstract

The global incidence of diabetes mellitus is on the rise, posing a significant health challenge worldwide. Recent evidence indicates a possible link between diabetes mellitus and temperature. This study aimed to examine the effect of temperature on diabetes patient visits in Yogyakarta Province, Indonesia, which is currently experiencing a high burden of diabetes and an increasingly aging population. Daily temperature data for 2021 were obtained from the Indonesian Meteorological, Climatological, and Geophysical Agency (BMKG). The number of diabetes patient visits was gathered from BPJS Health data samples for primary and referral healthcare facilities in the province. The relationship between temperature and diabetes patient visits was explored through data visualization, Pearson correlation, and Poisson regression models. We found a short-term correlation between higher temperatures and an increase in patient visits daily. However, observations on Sundays and holidays may not reflect the delayed effect of temperature on patient visits since not all primary and referral care providers offer services on these days. Nonetheless, including the observations on these days is necessary to assess the overall impact of temperature on patient visits weekly and monthly. The regression analysis revealed that for each additional 1°C increase in the average monthly temperature, the estimated number of patients for primary care increased by 15% and 14% for referral care services.

## Introduction

Diabetes mellitus (diabetes) is a chronic metabolic disorder that is characterized by high levels of glucose in the blood due to insufficient insulin production or the body’s inability to use insulin effectively. Symptoms of diabetes include frequent urination, excessive thirst, hunger, fatigue, blurred vision, and slow healing of cuts and bruises.^1^ Diabetes has a multifactorial etiology, which includes the interaction between genetic, lifestyle, and environmental factors. Several studies have highlighted the crucial role of lifestyle factors such as physical inactivity, unhealthy dietary habits, and obesity in the onset and progression of diabetes.^2^

As of 2021, an estimated 537 million adults between the ages of 20-79 are living with diabetes worldwide. This translates to one in every ten individuals within this age group. Projections indicate that by 2030, the number of adults living with diabetes will rise to 643 million; by 2045, it will increase to 783 million. In 2021, diabetes caused 6.7 million deaths globally, which amounts to one death every five seconds. Diabetes is also a significant economic burden, with healthcare expenditures caused by the condition exceeding at least USD 966 billion dollars, representing a 316% increase over the last 15 years.^3^ The majority of people affected by diabetes, over three-quarters, reside in low- and middle-income countries where the health systems may not have the resources to sufficiently address the issue.

In 2021, Indonesia had an estimated 19.5 million adults aged 20-79 years with diabetes, making it the fifth-highest country in the world with diabetes. This number is also expected to increase to 28.6 million in 2045.^3^ Most people with diabetes in Indonesia are diagnosed too late, often when they are already experiencing complications such as neuropathy, kidney failure or vision problems. Patients often seek medical attention when their diabetes has already progressed to an advanced stage, which can lead to serious health issues.^4^ The average annual cost of diabetes treatment in Indonesia is US 1,207.8 dollars per patient, with direct costs, especially for medicine and general practitioners’ visits, contributing the most to the overall cost.^5^ This amount is quite significant, considering that Indonesia’s per capita income in 2021 was around US 4,349.5 dollars.^6^ The high cost of diabetes treatment compared to the population’s average income highlights the financial burden and challenges that individuals with diabetes face in Indonesia.

A growing body of evidence suggests that weather conditions contribute to diabetes risk. Short-term exposure to extreme temperatures - whether it be hot or cold - can affect the morbidity and mortality rates of individuals with diabetes.^7,8^ The human body’s thermoregulatory function helps to maintain a stable core temperature regardless of external temperatures. It cools down through sweating and vasodilation in hot environments and increases temperature through metabolic heat production and vasoconstriction in cold environments.^9^ Diabetes patients have compromised abilities to dissipate heat and are therefore at a higher risk of heat-related illnesses. Due to reduced skin blood flow and sweating response during heat exposure, diabetes patients’ glycemic control can be adversely affected.^10^ Moreover, the elderly are particularly vulnerable to the negative impacts of heat exposure on diabetes.^7,11^ Raising awareness about the risks of temperatures and their impact on diabetes could potentially help reduce the burden of diabetes-related healthcare costs, e.g., by prioritizing the needs of vulnerable populations, such as the elderly, and incorporating education on managing diabetes in different weather conditions.

Indonesia is a country with a tropical climate and an average monthly temperature of 25°C-26°C.^12^ According to climate model projections, Indonesia is expected to be highly susceptible to severe heatwaves, and by the year 2080, it is anticipated to encounter exceedingly hazardous conditions nearly every day of the year under RCP8.5.^12^ Furthermore, with global temperatures set to reach record highs in the next five years (2023-2027),^13^ due to heat-trapping greenhouse gases and an upcoming El Niño event, it is crucial to gain a deeper understanding of the health consequences of extreme temperatures in tropical regions.

The Indonesian National Health Insurance scheme JKN (INHI) is a program launched by the Indonesian government in 2014 to provide equitable and comprehensive health services for all Indonesian citizens, and the mandate as the organizer of the JKN is carried out by The National Healthcare and Social Security Agency (Badan Penyelenggara Jaminan Sosial Kesehatan – BPJS Health).^14^ Each member of the insurance program is registered to a specific primary healthcare facility that acts as the initial point of contact for accessing the insurance system. Access to higher-level healthcare facilities and specialists (referral healthcare) is only permitted after first consulting with a primary healthcare facility registered with BPJS Health. The initial point of contact for primary healthcare can be a Puskesmas (public health center), a Klinik Pratama (primary clinic), or a general practitioner.^15^

BPJS Health provides sample data for research purposes that contains information on membership and healthcare services. This data records the utilization of BPJS Health services by its members at healthcare facilities that cooperate with BPJS Health and have submitted claims to BPJS Health. The sample data represents the entire membership and healthcare services data, which has been standardized and extracted to maintain data quality.^14^ The sample data provided by BPJS Health has been used for various research purposes, such as analyzing patterns of chronic disease multimorbidity,^16^ evaluating the dengue surveillance system,^17^ and investigating the relationship between temperature and primary healthcare visits. ^15^

A previous study using BPJS Health data linked with daily-district meteorological data investigated the relationship between temperature and primary healthcare visits. The study found that on hot days, all-cause visits increased by 8%, visits with a diabetes diagnosis increased by 25%, and visits with a cardiovascular disease diagnosis increased by 14%.^15^ In contrast, our study aims to explore the relationship between temperature and healthcare visits of patients with diabetes, not only at primary care but also referral care. The third edition of BPJS Health sample data (2015-2020), launched in December 2021, consists of general sample data and contextual sample data for Diabetes mellitus and Tuberculosis. Although both types of sample data contain the same variables, they differ in their methods of sample selection. Our study analyzes a novel dataset of contextual BPJS Health sample data for Diabetes mellitus and investigates its associations with environmental variables. Additionally, our study aims to investigate the delayed effect of temperature on healthcare visits by patients with diabetes.

## Results

The present study investigates the potential influence of ambient temperature on visits made by diabetes patients to primary and referral healthcare services in Yogyakarta province, Indonesia. Daily data on temperature and the number of patient visits were analyzed, and a lag time of up to four weeks was considered.

The results show that patient visits to healthcare services were higher from Monday to Saturday as compared to Sunday. On average, the number of visits for diabetes patients was highest on Mondays, and lowest on Sundays, for both primary and referral healthcare services. Fridays had the lowest average number of visits among weekdays (Figure 1A and 1B).

**Figure 1.**
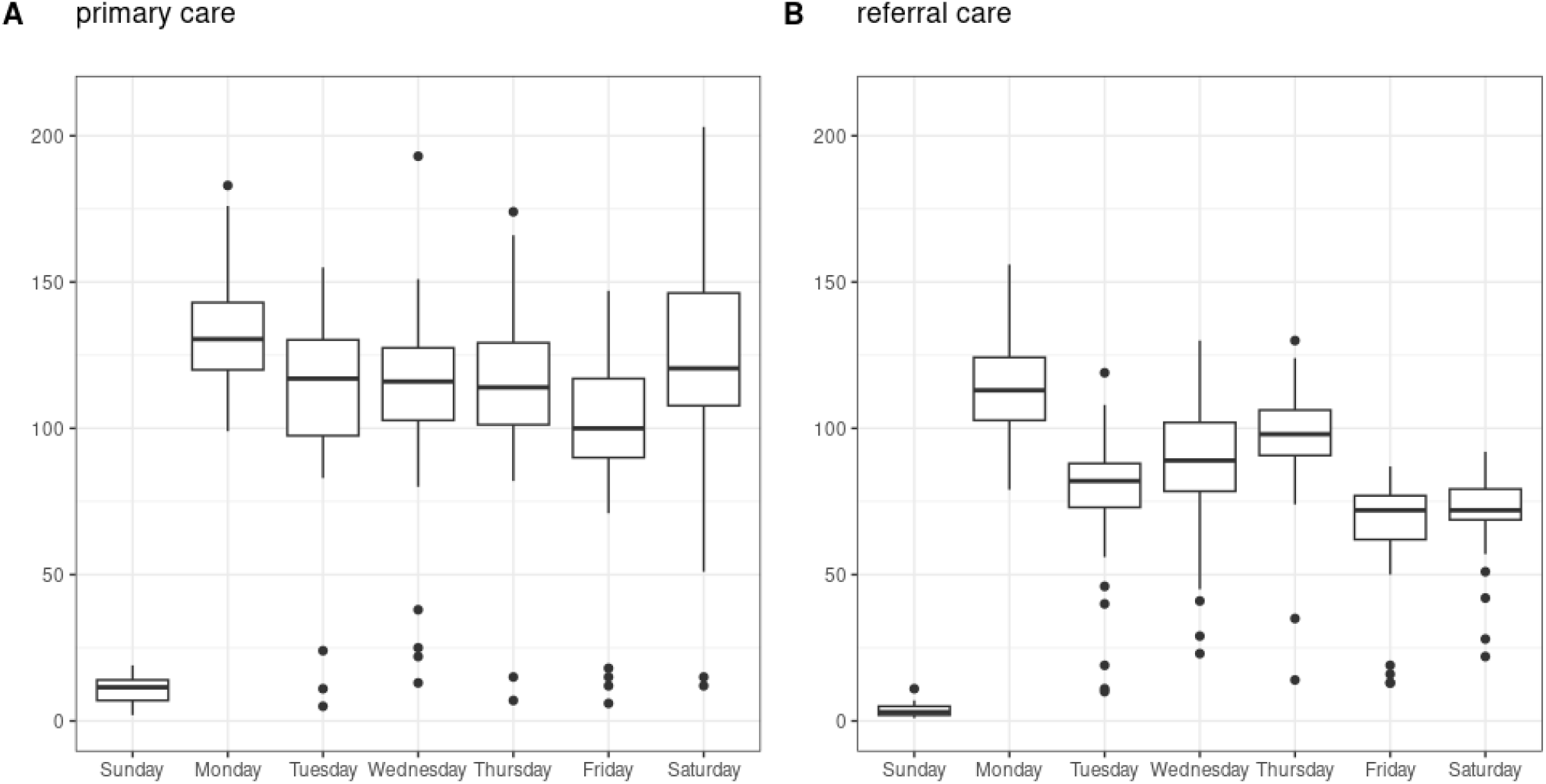
Pattern of the daily number of diabetes patients visiting primary and referral care services in a week

Our study revealed that many outliers with unusually low values were present from Tuesday through Saturday (Figure 1). To investigate this trend further, we analyzed the dataset considering the holidays occurring during the study period. The holidays were defined based on Joint Decree (SKB) No. 642/2020, No. 4/2020, and No. 4/2020, signed by the Minister of Religion, Minister of Manpower, and Minister of Administrative Reform and Bureaucratic Reform, Republic of Indonesia, on September 10, 2020. This analysis aimed to determine whether Sundays and holidays had an influence on the number of diabetes patient visits, and our findings suggest that Sundays and holidays had a consistently low number of diabetes patient visits (Figure 2 A-D). However, when the data was aggregated on a weekly basis, the number of weekly visits generally showed a similar pattern, regardless of whether Sundays and holidays were considered or not (Figures 2 E-H).

**Figure 2.**
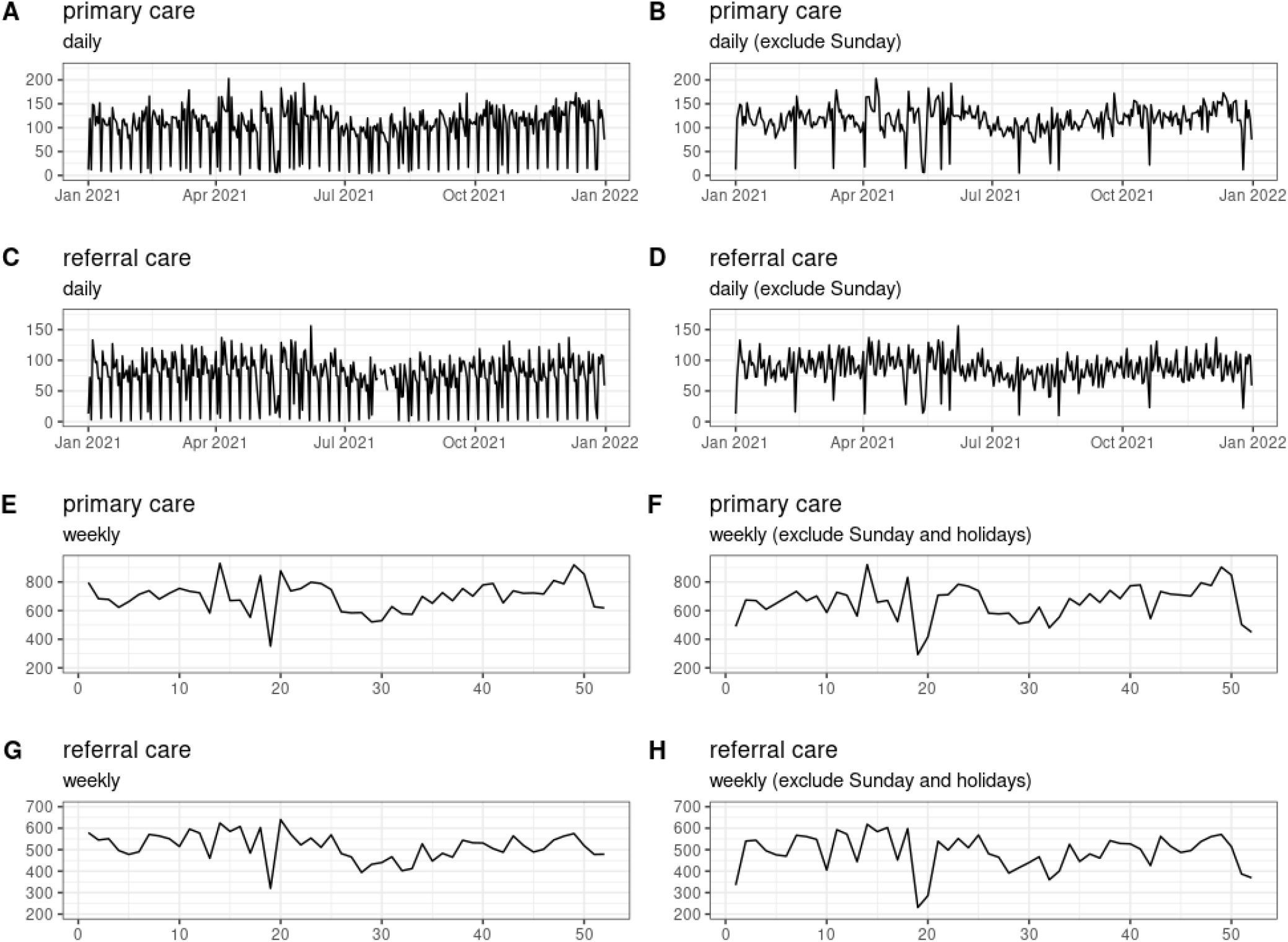
Time series graphs of the daily and weekly number of diabetes patients visiting primary and referral care services.

The study period in 2021 recorded temperatures ranging from 23.8°C to 28.8°C (Figure 3A). The temperature gradually increased from January to May, reaching its peak during the latter month. It then displayed a declining trend, hitting its lowest point around July-August, before rising again to its second peak in October. After October, the trend remained relatively flat until December (Figure 3 A-B). The trends in monthly diabetes patient visits to primary and referral care services mirrored those of the temperature data (Figure 3 C-F). In general, the number of patient visits to primary care was higher than the referral care.

**Figure 3.**
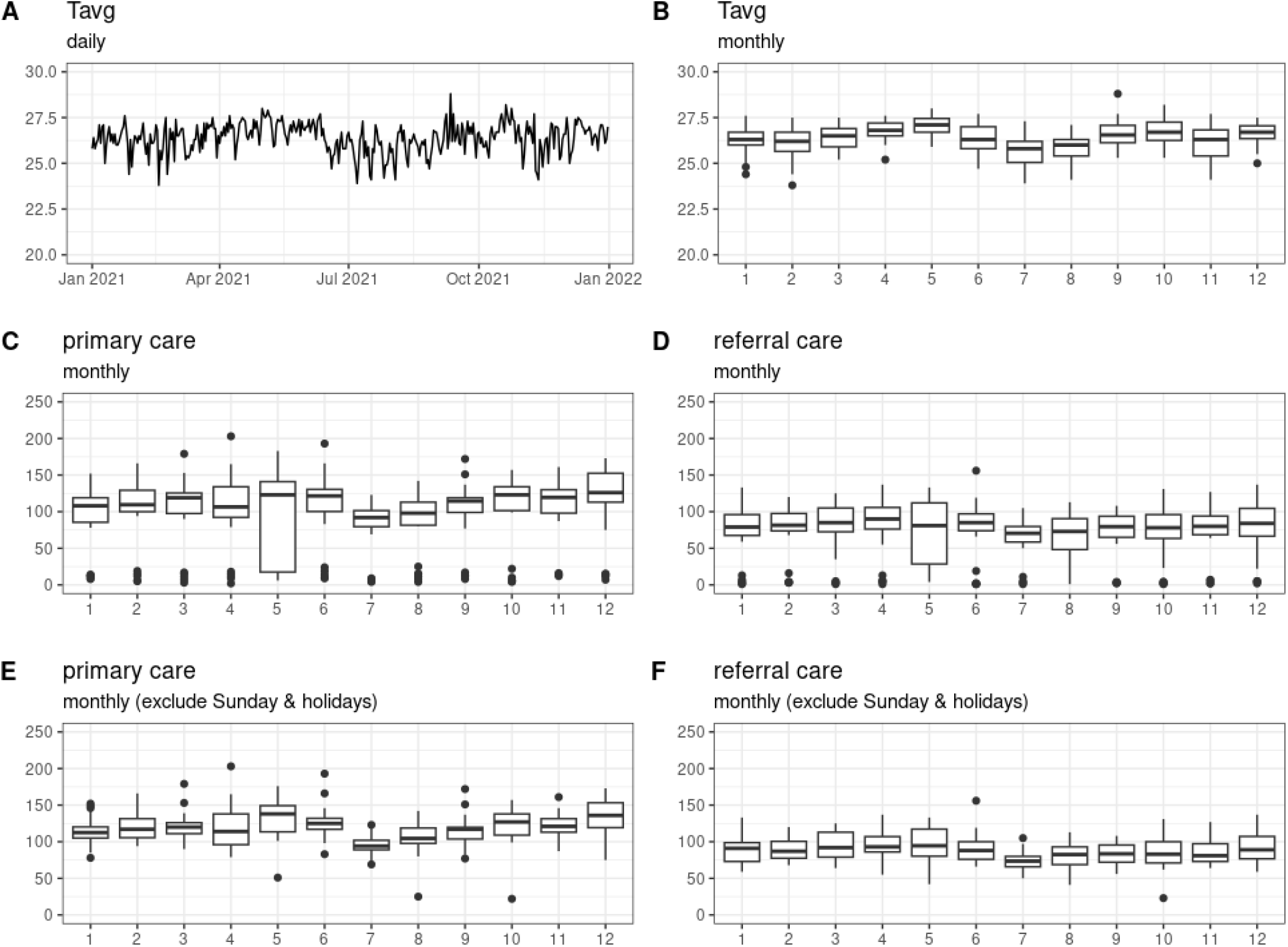
Time series graphs of the temperature and the monthly number of diabetes patients visiting primary and referral care services.

The Pearson correlation coefficient between the daily data for diabetes patient visits and temperature was found to be low, with a range of values from -0.023 (p-value > 0.05) at the 8-day lag to 0.109 (p-value < 0.05) at the 4-day lag for primary care (Figure 4A). For referral care, the range of Pearson correlation coefficient was from -0.035 (p-value > 0.05) at the 7-day lag to 0.133 (p-value < 0.05) at the 4-day lag (Figure 4B). The exclusion of observations on Sundays and holidays led to a consistently positive correlation coefficient. For primary care, this ranged from 0.073 (p-value > 0.05) at the 13-day lag to 0.248 (p-value < 0.05) at the 2-day lag (Figure 4C), while for referral care, the range was from 0.05 (p-value > 0.05) at the 13-day lag to 0.232 (p-value < 0.05) at the 11-day lag (Figure 4D). Aggregating the data on a weekly basis resulted in the highest and significant Pearson correlation coefficient occurring at a 2-week lag for both primary and referral care (Figure 4 E-F). However, when observations on Sundays and holidays were excluded and the data was aggregated weekly, there was no significant correlation (Figure 4 G-H).

**Figure 4.**
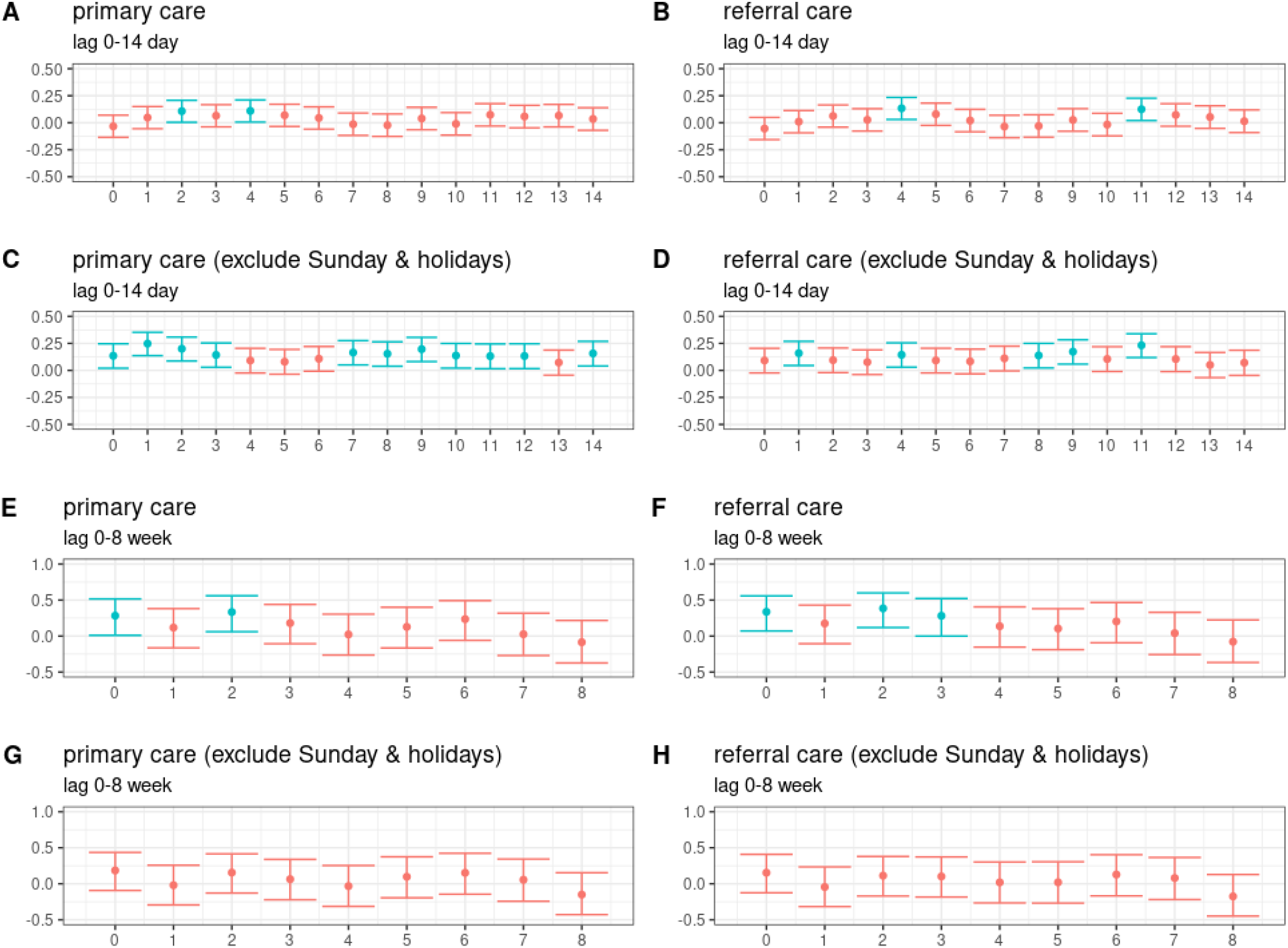
The Pearson correlation coefficient between temperature and the number of diabetes patients visits. *Red represents p-value > 0*.*05, indicating a non-significant result, and blue represents p-value < 0*.*05, indicating a significant result*.

The similar patterns observed in the increase and decrease trends of cumulative monthly visits and monthly average temperatures (Figure 3) prompted further investigation. For this, we developed a Poisson regression model to model the cumulative number of visits made by diabetes patients that is a rate based on the average monthly temperature. The results of the model suggest a positive correlation between the temperature and the number of patients visiting primary and referral care services. Our findings indicate for each additional 1°C increase in the average monthly temperature, there is an estimated 15% increase in the number of patients for primary care and 14% increase for referral care services (Figure 5).

**Figure 5.**
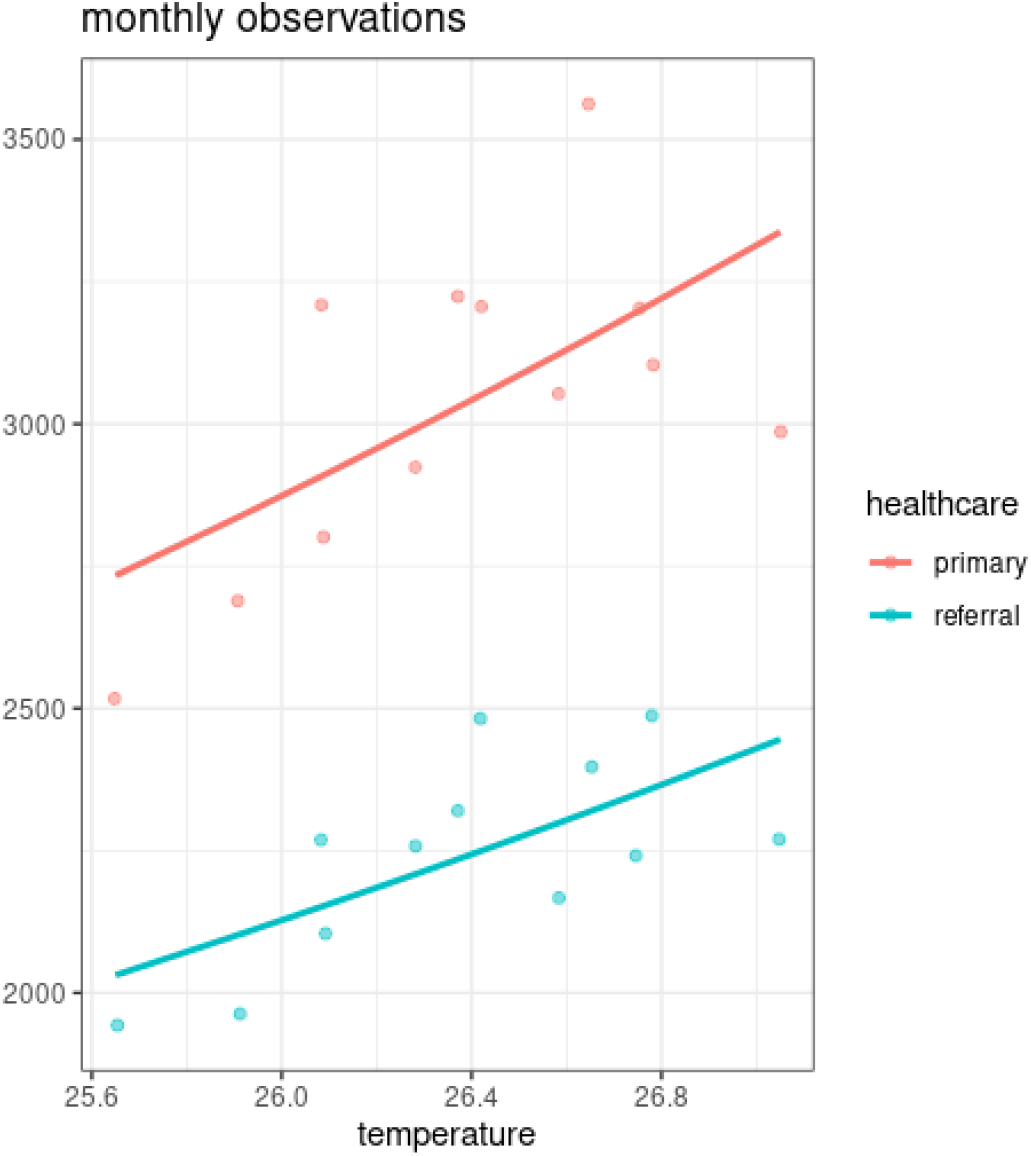
Temperature and the expected cumulative number of diabetes patients visits.

## Discussion

Studies investigating the effects of heat and cold on human health in tropical countries that experience small diurnal temperature ranges or minimal differences between hot and cold seasons are scarce.^18^ This gap in research hinders our understanding of the potential health consequences associated with exposure to extreme temperatures in these regions.

People generally feel comfortable in environments with temperatures ranging from 22°C to 27°C, depending on the humidity levels.^19^ However, during the study period in 2021, temperatures in our study area ranged from 23.8°C to 28.8°C (as shown in Figure 3), indicating that people in this area may have experienced uncomfortable thermal exposure at times, particularly due to heat. This is a concern, especially for people with diabetes, whose ability to effectively regulate their body temperature is compromised, ^10^ making them more susceptible to heat-related illnesses.

Our research has revealed a positive correlation between ambient temperature and the number of diabetes patients visiting primary and referral healthcare services. This suggests that as temperatures increase, the number of patient visits is likely to increase, particularly in the near future.^13^ We observed a short-term association in daily observations where there were many significant and positive correlations between higher temperatures and the number of patient visits, when the observations on Sundays and holidays were excluded (Figure 4 C-D). This relationship is particularly pronounced for visits to primary care (Figure 4C). It is important to note that the association between ambient temperature and patient visits does not hold true for Sundays and holidays. As many healthcare providers do not provide services on these days, very few diabetes patients visit healthcare services. Therefore, daily data collected on Sundays and holidays may not accurately represent the actual influence of temperature on patient visits (Figure 4 A-B).

However, the inclusion of the observations on Sundays and holidays are still essential when assessing weekly association of temperature and patients visit. Although a positive correlation trend is evident, excluding observations on Sundays and holidays produces no significant associations (Figures 4 G-H). This suggests that while not all primary and referral care providers offer services on Sundays and holidays, their inclusion is still important. People who experience adverse medical conditions, including those related to higher temperature exposure, may still seek emergency services on Sundays and holidays. Therefore, their inclusion in the data is still important for assessing the overall impact of temperature on patient visits.

The analysis shows a significant positive correlation for primary care in the week coinciding with higher temperatures and the second week following. The correlation is also seen in the second and third weeks following the temperature increase for referral care. These findings suggest that diabetes patients may require multiple visits to healthcare providers for adequate control of their condition. The observed lag in the correlation between temperature and patient visits may be due to several factors. For example, patients may delay seeking medical care until their symptoms become more severe or persistent. In addition, our findings suggest that follow-up visits may be necessary for effective management of diabetes during periods of temperature fluctuations.

The immediate impact of rising temperatures on the increased number of visits by diabetes patients to healthcare services was also pronounced when the analysis was carried out monthly. It was found that the pattern of increase and decrease in temperature and the number of visits looked similar, for both primary and referral care, regardless of whether considering visits on Sundays and holidays (Figure 3 C-F). In addition, using the Poisson regression model, the positive correlation was significant for primary and referral care (Appendix 2). Our findings highlight the need for healthcare providers to be aware of the potential increase in patient visits during hotter months and to prepare accordingly. To ensure that patients receive prompt and adequate care, it may be necessary to boost staffing levels, allocate more resources, and enhance outreach efforts. Additionally, health promotion initiatives should be implemented to raise patient awareness on managing diabetes during periods of elevated heat, which could involve staying hydrated, monitoring blood glucose levels more frequently, remaining indoors or in air-conditioned environments during the hottest times, and taking measures to protect medical supplies from excessive heat.^20^

Our study only included visits to healthcare facilities that have collaborated with BPJS Health and, therefore, may not reflect the overall prevalence of diabetes in the population. Furthermore, our analyses are based on the BPJS Health sample dataset that only covers 8.40% of the INHI members population in Yogyakarta province.^14^ However, despite these limitations, our findings are consistent with previous studies that have reported a positive association between higher temperature and diabetes-related health outcomes. Individuals who have diabetes are at a higher risk of experiencing negative impacts from heat exposure compared to those who do not have diabetes. ^20^ These negative impacts may include increased hospitalizations and emergency department visits due to diabetes-related complications. ^21,22^

The high prevalence of undiagnosed diabetes in low- and middle-income countries suggests that many individuals may not have access to appropriate medical care and may not be aware of their condition.^4,23^ This can pose a significant challenge in assessing the relationship between temperature and diabetes-related health outcomes in these regions. As the burden of diabetes is high in Yogyakarta^24^ and the region is experiencing a trend of a rising aging population^25^, it is important to take proactive measures to mitigate the potential adverse health effects. Our study highlights the critical role of regional health policy in mitigating the impact of temperature on diabetes patients and reducing the overall burden of the disease on healthcare systems. The regional health office should take proactive steps to raise public awareness about the potential risks of temperature exposure to diabetes patients. Moreover, as older adults with diabetes are particularly vulnerable to the negative health effects of temperature, the regional health office should develop targeted outreach programs that provide additional support for this population.^26^ This may include home visits, educational materials, and assistance with accessing healthcare services during periods of high temperature. By addressing these issues, healthcare systems can better respond to the potential health consequences of extreme temperature events and help prevent adverse health outcomes among vulnerable populations.

## Methods

### Design and setting

We conducted population-based study to investigate the relationship between ambient temperature and visits to healthcare facilities by diabetes patients in Yogyakarta Province, Indonesia. Yogyakarta is a province located in the central southern part of Java Island, approximately 450 kilometers southeast of the Jakarta Capital Region. According to the 2010 and 2020 Census data, the population of Yogyakarta Province has increased from 3,457,491 to 3,668,719 people. As of mid-2022, the estimated population had further grown to 3,761,870. Despite being the second smallest province in Indonesia, with an area of 3,170.65 km^2^, it has one of the highest population densities on Java Island, as well as in the surrounding areas of Central Java.^27^ Yogyakarta Province has a tropical climate classified as Am under the Köppen-Geiger system. This classification indicates a tropical monsoon climate with a short dry season. Yogyakarta Province’s climate is characterized by warm temperatures, high humidity, and a distinct wet and dry season, with a shorter dry season than many other regions with a tropical monsoon climate.^28^ According to the 2018 Basic Health Research (Riskesdas) report, Yogyakarta Province ranks third among Indonesian provinces with the highest prevalence of diabetes, with a prevalence rate of 3.1%. Moreover, BPJS Health claims for Diabetes mellitus from 2015-2019 have increased rapidly in Yogyakarta Province, indicating a growing burden of the disease. This highlights the need for effective prevention and management strategies in primary care to prevent diabetes consulting services or internal medicine specialists in referral care services from being overwhelmed.^24^

### Study Population and Visitation Data

The study used the third edition of BPJS Health sample data, which was launched in December 2021 and covers the years 2015 to 2021. The dataset includes both general and contextual sample data, with the latter including diabetes mellitus and tuberculosis. Although both types of sample data have the same variables, they differ in their methods of sample selection. The contextual sample data for diabetes mellitus is derived from membership and service data from INHI members through the BPJS Health data warehouse. The study selected participants who had received primary or referral care services with a diabetes diagnosis in 2019, based on ICD-10 codes E10 (Type 1 diabetes mellitus) and E11 (Type 2 diabetes mellitus without complications). Service data for these participants were collected in 2021, including primary and/or referral care utilization. Participants who used both primary and referral care services were not unique, meaning they could have made more than one visit to each service. Yogyakarta province has a population of 44,696 INHI members and a sample size of 3,760 contextual diabetes mellitus participant, which is 8.40% of the INHI members population. The margin of error is 1.50%, which means that the survey results are within 1.50 percentage points of the true population value 95% of the time.^14^

### Temperature Data

We obtained daily temperature data from the Sleman Geophysics Monitoring Station, operated by the Indonesian Meteorology, Climatology, and Geophysics Agency (BMKG). This station, which has the World Meteorological Organization (WMO) ID 96855, is located at coordinates -7.82000 latitude and 110.30000 longitude. The data included daily average temperatures (°C) from January 1st to December 31st, 2021.^29^ To analyze the association between temperature and diabetes mellitus patients’ visits to healthcare facilities, we estimated weekly and monthly average temperatures. We also explored the lagged effects of temperature variables on diabetes mellitus patients’ visits to healthcare facilities to investigate the delayed effects of exposure on outcomes. We accounted for possible lag effects by including lag times of up to 14 days and 4 weeks in our analysis, encompassing lag 1–14 days and lag 1–4 weeks of the temperature variable, respectively.

### Analysis

In the first stage of our analysis, we employed data visualization techniques to explore the relationship between temperature and the number of patient visits. We utilized line plots and box plots to visualize the data at different temporal scales, including daily, weekly, and monthly. This allowed us to identify patterns and trends in the relationship between variables, such as seasonal variations, that could vary depending on the level of aggregation. In the second stage, we calculated the Pearson correlation coefficients to quantify the strength and direction of the relationship between temperature and the number of visits. Specifically, we assessed the delayed effect of temperature on patients’ visits by considering up to a 14-day lag for the daily temporal scale and up to a 4-week lag for the weekly temporal scale. We estimated the Pearson correlation coefficients and their confidence intervals and visualized them to identify any delay effects. Finally, we utilized Poisson regression to model the cumulative number of DM patients visiting monthly. We calculated the cumulative number of diabetes patients visiting monthly by summing the daily visit counts for each month and used this variable as our response variable. We used the mean temperature for each month as our predictor variable. This Poisson regression model allowed us to estimate the effect of temperature on the cumulative number of diabetes patients visiting monthly and gain insights into the potential impact of temperature on primary and referral healthcare services. All analyses were performed in R,^30^ and the code needed to reproduce the results presented here is available on GitHub at https://github.com/alramadona/DM-temp.

### Ethical Considerations

The study relied on secondary data from BMKG and BPJS Health and did not require ethical approval since it did not involve human participants. BPJS Health granted the author permission to use their anonymized individual health insurance data sample.

### Data Availability

The data supporting the findings of this study are available online from the BPJS Health and BMKG data portals at https://data.bpjs-kesehatan.go.id/ and http://dataonline.bmkg.go.id/,respectively.

## Acknowledgements (not compulsory)

We wish to express our sincere gratitude to BPJS Health and BMKG for granting us access to their valuable dataset, which formed the basis of our research.

## Author contributions statement

AR, HK, and LL designed the study and wrote the manuscript. AR and HK conceptualized the experiments, while AR conducted them. AR, LL, and AF collected data and managed the database. AR, HK, and LL analyzed the results, and all authors discussed the findings, reviewed, and approved the final manuscript.

