## Supplementary Information for "Effect of Ambient Temperature on Diabetes Patient Visits at Primary and Referral Healthcare Services in Yogyakarta Province, Indonesia: An Analysis of Indonesian National Health Insurance Data 2021"

### Suppl. Table. 1. Pearson Correlation

| **Daily data** | | | | | | |  | **Daily data (excluding Sundays and holidays)** | | | | | | |
| --- | --- | --- | --- | --- | --- | --- | --- | --- | --- | --- | --- | --- | --- | --- |
| **healthcare** | **lag** | **estimate** | **lower** | **upper** | **p.value** | **sig.** |  | **healthcare** | **lag** | **estimate** | **lower** | **upper** | **p.value** | **sig.** |
| primary care | 0 | -0.033 | -0.135 | 0.07 | 0.53 |  |  | primary care | 0 | 0.135 | 0.021 | 0.246 | 0.021 | * |
| primary care | 1 | 0.048 | -0.055 | 0.15 | 0.365 |  |  | primary care | 1 | 0.248 | 0.137 | 0.353 | 0 | * |
| primary care | 2 | 0.107 | 0.004 | 0.208 | 0.041 | * |  | primary care | 2 | 0.2 | 0.087 | 0.308 | 0.001 | * |
| primary care | 3 | 0.065 | -0.038 | 0.167 | 0.218 |  |  | primary care | 3 | 0.143 | 0.029 | 0.254 | 0.015 | * |
| primary care | 4 | 0.109 | 0.006 | 0.21 | 0.038 | * |  | primary care | 4 | 0.091 | -0.024 | 0.205 | 0.12 |  |
| primary care | 5 | 0.069 | -0.035 | 0.171 | 0.191 |  |  | primary care | 5 | 0.08 | -0.035 | 0.194 | 0.173 |  |
| primary care | 6 | 0.044 | -0.06 | 0.147 | 0.404 |  |  | primary care | 6 | 0.108 | -0.008 | 0.221 | 0.067 |  |
| primary care | 7 | -0.014 | -0.118 | 0.09 | 0.789 |  |  | primary care | 7 | 0.165 | 0.051 | 0.276 | 0.005 | * |
| primary care | 8 | -0.023 | -0.126 | 0.081 | 0.666 |  |  | primary care | 8 | 0.154 | 0.039 | 0.265 | 0.009 | * |
| primary care | 9 | 0.039 | -0.065 | 0.143 | 0.459 |  |  | primary care | 9 | 0.196 | 0.082 | 0.306 | 0.001 | * |
| primary care | 10 | -0.011 | -0.115 | 0.093 | 0.84 |  |  | primary care | 10 | 0.137 | 0.021 | 0.249 | 0.021 | * |
| primary care | 11 | 0.074 | -0.031 | 0.176 | 0.167 |  |  | primary care | 11 | 0.132 | 0.016 | 0.245 | 0.026 | * |
| primary care | 12 | 0.058 | -0.047 | 0.161 | 0.281 |  |  | primary care | 12 | 0.133 | 0.017 | 0.246 | 0.025 | * |
| primary care | 13 | 0.066 | -0.039 | 0.17 | 0.216 |  |  | primary care | 13 | 0.073 | -0.044 | 0.188 | 0.223 |  |
| primary care | 14 | 0.035 | -0.07 | 0.139 | 0.514 |  |  | primary care | 14 | 0.157 | 0.041 | 0.269 | 0.008 | * |
| referral care | 0 | -0.054 | -0.156 | 0.049 | 0.304 |  |  | referral care | 0 | 0.092 | -0.024 | 0.204 | 0.119 |  |
| referral care | 1 | 0.01 | -0.094 | 0.113 | 0.855 |  |  | referral care | 1 | 0.159 | 0.045 | 0.269 | 0.006 | * |
| referral care | 2 | 0.062 | -0.041 | 0.164 | 0.238 |  |  | referral care | 2 | 0.095 | -0.02 | 0.208 | 0.106 |  |
| referral care | 3 | 0.027 | -0.077 | 0.13 | 0.615 |  |  | referral care | 3 | 0.077 | -0.039 | 0.19 | 0.192 |  |
| referral care | 4 | 0.133 | 0.03 | 0.234 | 0.011 | * |  | referral care | 4 | 0.144 | 0.03 | 0.255 | 0.014 | * |
| referral care | 5 | 0.079 | -0.025 | 0.181 | 0.134 |  |  | referral care | 5 | 0.092 | -0.024 | 0.205 | 0.118 |  |
| referral care | 6 | 0.02 | -0.084 | 0.124 | 0.702 |  |  | referral care | 6 | 0.083 | -0.032 | 0.197 | 0.158 |  |
| referral care | 7 | -0.035 | -0.138 | 0.069 | 0.51 |  |  | referral care | 7 | 0.111 | -0.004 | 0.224 | 0.06 |  |
| referral care | 8 | -0.03 | -0.134 | 0.074 | 0.568 |  |  | referral care | 8 | 0.138 | 0.023 | 0.25 | 0.019 | * |
| referral care | 9 | 0.026 | -0.079 | 0.13 | 0.627 |  |  | referral care | 9 | 0.173 | 0.058 | 0.283 | 0.003 | * |
| referral care | 10 | -0.017 | -0.122 | 0.087 | 0.744 |  |  | referral care | 10 | 0.106 | -0.011 | 0.219 | 0.075 |  |
| referral care | 11 | 0.125 | 0.02 | 0.226 | 0.019 | * |  | referral care | 11 | 0.232 | 0.119 | 0.339 | 0 | * |
| referral care | 12 | 0.073 | -0.032 | 0.176 | 0.175 |  |  | referral care | 12 | 0.105 | -0.011 | 0.219 | 0.077 |  |
| referral care | 13 | 0.053 | -0.052 | 0.157 | 0.323 |  |  | referral care | 13 | 0.05 | -0.067 | 0.166 | 0.399 |  |
| referral care | 14 | 0.014 | -0.091 | 0.119 | 0.794 |  |  | referral care | 14 | 0.071 | -0.046 | 0.187 | 0.235 |  |

| **Weekly data** | | | | | | |  | **Weekly data (excluding Sundays and holidays)** | | | | | | |
| --- | --- | --- | --- | --- | --- | --- | --- | --- | --- | --- | --- | --- | --- | --- |
| **healthcare** | **lag** | **estimate** | **lower** | **upper** | **p.value** | **sig.** |  | **healthcare** | **lag** | **estimate** | **lower** | **upper** | **p.value** | **sig.** |
| primary care | 0 | 0.283 | 0.011 | 0.516 | 0.042 | * |  | primary care | 0 | 0.184 | -0.093 | 0.435 | 0.191 |  |
| primary care | 1 | 0.118 | -0.163 | 0.381 | 0.411 |  |  | primary care | 1 | -0.02 | -0.294 | 0.257 | 0.888 |  |
| primary care | 2 | 0.334 | 0.061 | 0.56 | 0.018 | * |  | primary care | 2 | 0.155 | -0.129 | 0.415 | 0.283 |  |
| primary care | 3 | 0.18 | -0.106 | 0.439 | 0.215 |  |  | primary care | 3 | 0.063 | -0.222 | 0.338 | 0.667 |  |
| primary care | 4 | 0.023 | -0.263 | 0.305 | 0.879 |  |  | primary care | 4 | -0.033 | -0.314 | 0.254 | 0.826 |  |
| primary care | 5 | 0.128 | -0.165 | 0.4 | 0.391 |  |  | primary care | 5 | 0.097 | -0.195 | 0.374 | 0.515 |  |
| primary care | 6 | 0.235 | -0.059 | 0.492 | 0.115 |  |  | primary care | 6 | 0.152 | -0.145 | 0.423 | 0.315 |  |
| primary care | 7 | 0.026 | -0.269 | 0.317 | 0.864 |  |  | primary care | 7 | 0.055 | -0.243 | 0.343 | 0.721 |  |
| primary care | 8 | -0.085 | -0.373 | 0.217 | 0.581 |  |  | primary care | 8 | -0.151 | -0.429 | 0.153 | 0.328 |  |
| referral care | 0 | 0.337 | 0.071 | 0.559 | 0.015 | * |  | referral care | 0 | 0.153 | -0.125 | 0.409 | 0.279 |  |
| referral care | 1 | 0.175 | -0.106 | 0.43 | 0.22 |  |  | referral care | 1 | -0.045 | -0.317 | 0.233 | 0.753 |  |
| referral care | 2 | 0.385 | 0.119 | 0.599 | 0.006 | * |  | referral care | 2 | 0.113 | -0.171 | 0.379 | 0.437 |  |
| referral care | 3 | 0.282 | 0 | 0.521 | 0.05 | * |  | referral care | 3 | 0.102 | -0.185 | 0.372 | 0.488 |  |
| referral care | 4 | 0.137 | -0.153 | 0.405 | 0.354 |  |  | referral care | 4 | 0.019 | -0.266 | 0.302 | 0.897 |  |
| referral care | 5 | 0.104 | -0.189 | 0.38 | 0.485 |  |  | referral care | 5 | 0.02 | -0.269 | 0.305 | 0.894 |  |
| referral care | 6 | 0.203 | -0.093 | 0.466 | 0.176 |  |  | referral care | 6 | 0.128 | -0.169 | 0.403 | 0.398 |  |
| referral care | 7 | 0.04 | -0.256 | 0.33 | 0.793 |  |  | referral care | 7 | 0.08 | -0.219 | 0.365 | 0.603 |  |
| referral care | 8 | -0.078 | -0.366 | 0.224 | 0.615 |  |  | referral care | 8 | -0.175 | -0.449 | 0.128 | 0.255 |  |

### Suppl. Table. 2. Poisson Regression Coefficients

|  |  | **estimate** | **lower** | **upper** | **p.value** |
| --- | --- | --- | --- | --- | --- |
| - primary care | (Intercept) | 4.257 | 1.142 | 7.371 | < 0.05 |
|  | temperature | 0.142 | 0.024 | 0.261 | < 0.05 |
| - referral care | (Intercept) | 4.216 | 2.035 | 6.395 | < 0.05 |
|  | temperature | 0.132 | 0.050 | 0.215 | < 0.05 |
